# Association between dopamine receptor D2 genetic polymorphism TaqIA1 (C t) and susceptibility to alcohol dependence

**DOI:** 10.1101/2020.04.15.20066498

**Authors:** Amrita Choudhary, Upendra Yadav, Pradeep Kumar, Vandana Rai

## Abstract

Several studies are published, which investigated dopamine receptor 2 (DRD2) gene TaqIA polymorphism as ris factor for alcohol dependence (AD) with positive and negative association. To derive a more precise estimation of the relationship, a meta-analysis of case-control studies that examined the association between DRD2 gene Taq1A polymorphism and alcohol dependence were performed. Eligible articles were identified through search of databases including PubMed, Science Direct, Springer link and Google Scholar. The association between the DRD2 TaqIA polymorphism and AD susceptibility was conducted using odds ratios (ORs) and 95 % confidence intervals (95 % CIs) as association measure.

A total of 69 studies with 9,125 cases and 9,123 healthy controls were included in current meta-analysis. Results of present analysis showed significant association between DRD2 TaqIA polymorphism and AD risk using a five genetic modes (allele contrast model -OR=1.22, 95% CI=1.13-1.32, p<0.0001; homozygote model -OR= 1.35, 95%CI= 1.18-1.55; p= <0.0001; dominant model -OR= 1.29; 95%CI= 1.20-1.39; p<0.0001; recessive model-OR= 1.21; 95%CI= 1.08-1.36; p= 0.0006). There was no significant association found between In subgroup analysis, TaqIA polymorphism was not significantly associated with AD risk in Asian population under all genetic models, but in Caucasian population TaqIA polymorphism was significantly associated with AD risk.

Overall, results support the hypothesis that DRD2 Taq1A polymorphism plays a role in alcohol dependence.

## Introduction

Alcoholism dependence (AD) is a complex behavioural and multifactorial disorder often associated with increased risk of developing other behavioural and psychiatric disorders such as anxiety, depression, anti social personality disorder and bipolar disorder, by affecting various neural mechanisms and part of brain with its effect according to the dose of consumption, genetic factors, etc. AD is a psychiatric disorder, with the life time population risk of approximately 5.4%.Family and twins studies support the role of a genetic component in AD. It is widely accepted that the dopamingeric system play a crucial role in the development of psychoactive substance dependence including opiates, cocaine, nicotine and alcohol (Koob and Moal, 2001 Munafò et al., 2001 Lingford-Hughes and Nutt,2003). The dopaminergic system regulates brain reward mechanism (Dick and Foroud,2003 Tupala and Tiihonen,2006), so genes of reward pathway especially dopamine receptors are considered a strong candidate for alcohol dependence. Alcohol stimulates dopamine receptors, which release dopamine in the ventral striatum leading to increased alcohol consumption through mechanisms involving incentive salience attributions and craving (Kienast and Heinz,2006). Aberrant dopamine signaling is implicated in several psychiatric and neurological brain disorders such as schizophrenia, depression, eating disorders, Parkinson’s disease and addiction (Glatt et al., 2003).

Dopamine D2 receptor (DRD2) gene is the most studies polymorphic site in the reward pathway in connection to AD. DRD2 receptor is present throughout the brain but the highest density was reported in substantia nigra, ventral tegmental area, and nucleus accumbens (Boyson et al., 1986Meador-Woodruff et al.,1989 Weiner and Brann,1989). DRD2 gene is present on chromosome 11q23.1 (Grandy et al. 1989 Eubanks et al.,1992), and after transcription it produces two transcripts that encode two isoforms (D2L isoform and D2S isoform). Several polymorphism are reported in DRD2 gene, but the most studied polymorphism is TaqIA polymorphism (rs1800497 C32806T Glu713Lys) in the 3 flanking region of the gene (Thompson et al.,1997). The *DRD2* gene spans approximately 270 kb with about a 250-kb intron separating the first and second exons (Eubanks et al., 1992). The TaqIA polymorphism lies 10,541 bp downstream (C32806T) of the termination codon of the DRD2 gene and fall within ankyrin repeat and kinase domain containing 1 (ANKK1) gene (Noble 2003 Neville et al.,2004). The two alleles are referred to as A2 (cytosine) and A1 (thymine) and the TaqIA genotypes are named A2/A2 (homozygous wid), A2/A1 (heterozygous) and A1/A1 (homozygous).DRD2 Taq1A polymorphism has been associated with reduced D2 receptor availability in the striatum (Thompson et al. 1997; Noble et al. 1991), lower mean relative glucose metabolic rate in dopaminergic regions (Noble et al. 1997), and low receptor density (Pohjalainen et al. 1998).

Blum et a (1990) initially demonstrated an association of the minor *Taq*l A allele (A1) of the DRD2 gene with AD. Since then, several studies have published from different populations, with many affirming this finding (Parsian et al. 1991; Amadeo et al. 1993; Noble et al. 1994 ; Neiswanger et al. 1995; Hietala et al. 1997; Lawford et al. 1997; Vasconceos et al.,2015; Panduro et al.,2017), while others have not (Geijer et al.,1994; Heinz et al.,1996; Sander et al. 1999; Kasiakogia-Worlley et al., 2011; Ragia et al.,2016). Hence, authors performed an update meta-analysis to validate the association between TaqIA polymorphism and alcohol dependence.

## Methods

Meta-analysis was carried out according to Meta-analysis of observational studies in epidemiology (MOOSE) guidelines (Stroup et al.,2000).

### Article Search

Published articles examining the effect of the DRD2 gene Taq1A polymorphisms on the risk of alcohol dependence were identified through electronic database searches in Pubmed, Science direct, Google scholar and springer link. Databases searches were done from January 1990 (the year that DRD2 Taq1A polymorphisms were first reported as a risk factor for AD. Electronic database searches were supplemented by manual searches of references of review and published research articles. The following terms were used: ‘‘Dopamine receptor 2’’, ‘‘DRD2,’’ ‘‘Taq1A’’, ‘‘Alcoholism’’, ‘‘Alcohol dependence’’, ‘‘polymorphism’’. Literature search included all languages. If any study reported results on different subpopulations according to ethnicity, each subpopulation was included as a separate study in the meta-analysis.

### Inclusion criteria

Studies included in our meta-analyses were based on the following criteria: (1) published should be published, (2) article should reported sample size, ancestry of samples etc ; and (3) sample not duplicative of other reports. Studies were excluded if: (1) incomplete raw data/information and not providing complete information for number of genotype and/or allele number calculation, (3) studies based on pedigree and (4) review, letter to editors and book chapters.

### Data Extraction

From each of the included articles the following information was extracted: first author family name, year of publication, country, ethnicity, study design, number of controls and cases, and evidence of HWE in controls.

### Statistical Analyses

Crude odds ratios (ORs) with 95% confidence intervals (CIs) were calculated under five genetic models: the allele model (mutant [M] allele vs. wild [W] allele), the dominant model (MM+WM vs. WW), the recessive model (MM vs. WM+WW), the homozygous model (MM vs. WW), and the heterozygous model (WM vs. WW). The OR was estimated by using fixed effects (Mantel and Haenszel,1959) and random effects (DerSimonian and Laird,1986) model depending upon heterogeneity. If higher heterogeneity between studies then the pooled OR is estimated using the RE model (Zintzaras and Ioannidis, 2004; Zintzaras et al., 2006). The heterogeneity was tested using the Q-statistic and was quantified using the I^2^ statistic (Higgins and Thompson,2002). Subgroup analysis based on ethnicity of subjects was performed to evaluate the stability of the results and sensitivity analysis was done by removing the studies not in Hardy–Weinberg equilibrium (HWE), and studies with small sample size. Control population of each study was tested for Hardy–Weinberg Equilibrium (HWE) using the chi square test.

Funnel plot and the Egger regression asymmetry test (Egger et al.,1997) were used for assessment of publication bias. P values were two-tailed with a significance level of 0.05. All analyses were done by the program Meta-Analyst (Wallace et al.,2013) and Mix version 1.7 (Bax et al.,2006).

## Results

### Literature Search

The flow chart of the study selection process is shown in Figure 1. Initial search of Pubmed, Science direct, Google Scholar and Springer Link databases, total 511 articles were retrieved, but 301 articles did not meet the inclusion criteria after reviewing abstract. Out of remaining 210 articles, 142 articles also excluded as not following inclusion criteria. After applying inclusion and exclusion criteria, total 68 studies were found suitable for the present meta-analysis (Blum et al., 1990; Bolos et al. 1990; Blum et al.,1991; Comings et al. 1991; Gelernter et al. 1991; Parsian et al. 1991; Schwab et al., 1991; Amadeo et al. 1993; Cook et al. 1992;Goldman et al. 1992; Arinami et al. 1993; Goldman et al. 1993; Comings et al. 1994; Geijer et al.,1994; Noble et al. 1994 ; Neiswanger et al. 1995; Sander et al. 1995; Chen et al. 1996; Finckh et al.,1996; Heinz et al.,1996; Lu et al. 1996; Chen et al. 1997; Goldman et al., 1997; Hietala et al. 1997; Kono et al. 1997; Lawford et al. 1997; Lee et al. 1997; Ishiguro et al. 1998; Gelernter and Kranzler 1999; Ovchinnikov et al. 1999; Sander et al. 1999; Amadeo et al. 2000; Bau et al. 2000; Gorwood et al. 2000; Samochowiec et al. 2000; Anghelescu et al. 2001; Lu et al., 2001; Matsushita et al. 2001; Pastorelli et al. 2001;Shaikh et al. 2001; Limosin et al. 2002; Foley et al. 2004; Konishi et al. 2004; Berggren et al. 2006;Freire et al. 2006; Huang et al. 2007; Sakai et al. 2007; Wang et al. 2007; Joe et al.2008; Namkoong et al. 2008; Ponce et al., 2008; Samochowiec et al., 2008; Wu et al., 2008; Kraschewski et al., 2009; Berggren et al.,2010; Bhaskar et al., 2010; Kovanen et al., 2010; Prasad et al.,2010; Kasiakogia-Worlley et al., 2011; Landgren et al., 2011; Mignini et al., 2012; Schellekens et al. 2012; Singh et al., 2013; Jasiewicz et al., 2014; Vasconceos et al.,2015; Ragia et al.,2016; Panduro et al.,2017). Panduro et al. (2017) analyzed samples from two population, so in present meta-analysis data of both the population were included as separate studies, hence total number of studies included in current meta-analysis was sixty nine.

**Figure 1.**
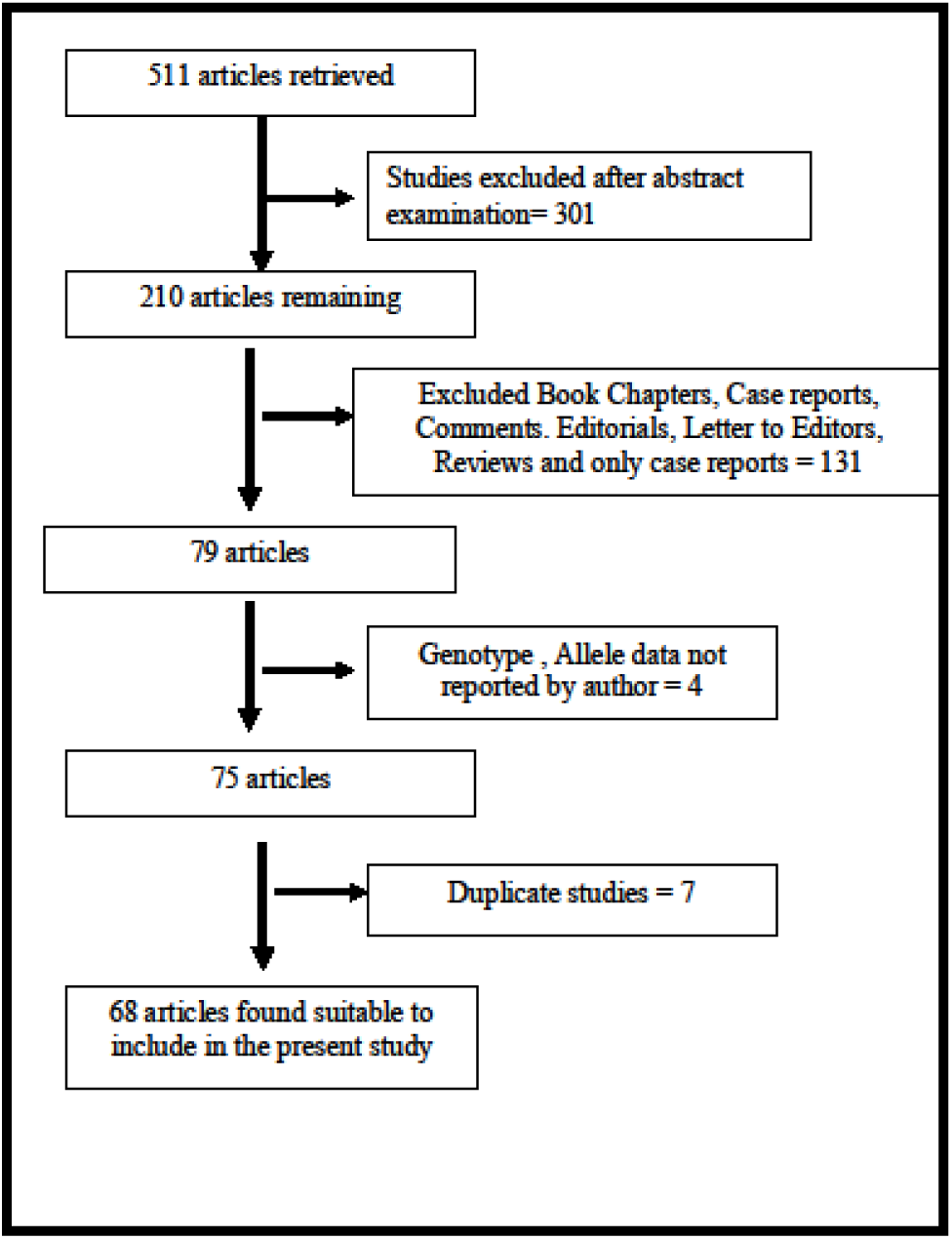
Study selection flow diagram.

### Characteristic of Eligible Studies

First study was published in 1990 (Blum et al., 1990) and recent study was published in 2017 (Panduro et al.,2017). In one study, authors mentioned only allele numbers. A total of 2591 subjects were involved in this meta-analysis, including 9125 AD cases and 9123 healthy controls. In cases, CC, CT and TT genotypes were 3903, 3063 and 1020 respectively. Out of 69 studies, control population of four studies (Bolos et al. 1990; Schwab et al., 1991; Kovanen et al.,2010; Panduro et al.,2017)are not in Hardy Weinberg equilibrium. Twenty of 69 studies were conducted in Asian populations, and the other 49 studies in Caucasian populations.

### Meta-analysis

Table 2 summarizes the ORs with corresponding 95% CIs for association between DRD2 TaqIA polymorphism and risk of alcohol dependence in allele contrast ((T vs. C/A1 vs. A2)), homozygote (TT vs. CC/A1A1 vs. A2A2), dominant (TT+CT vs. CC/A1A1+A1A2 vs. A2A2), recessive (TT vs. CT+CC/ A1A1 vs. A1A2 + A2A2) and co-dominant (CT vs. CC/A1A2 vs. A2A2) models.

**Table 1.**
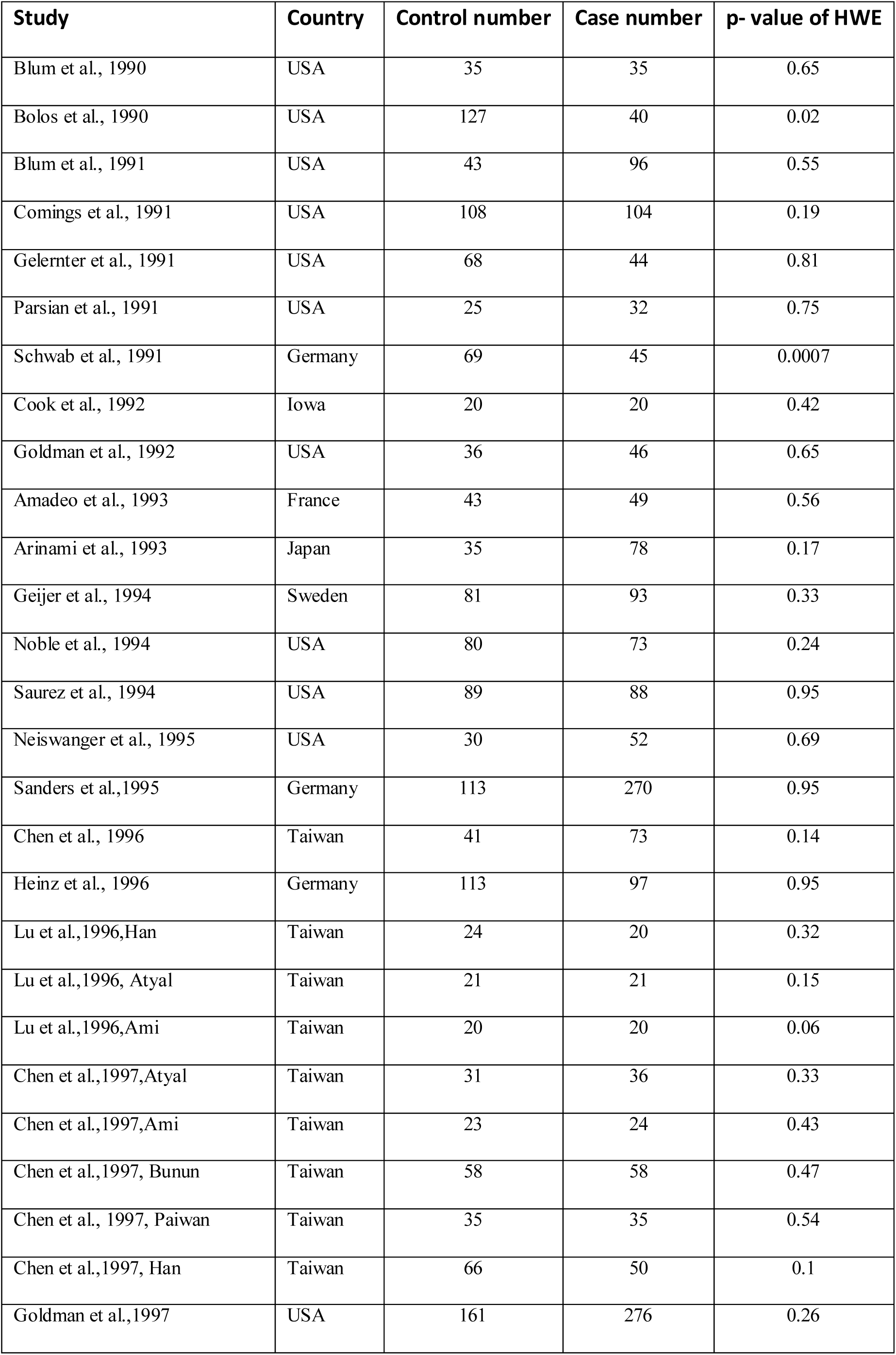
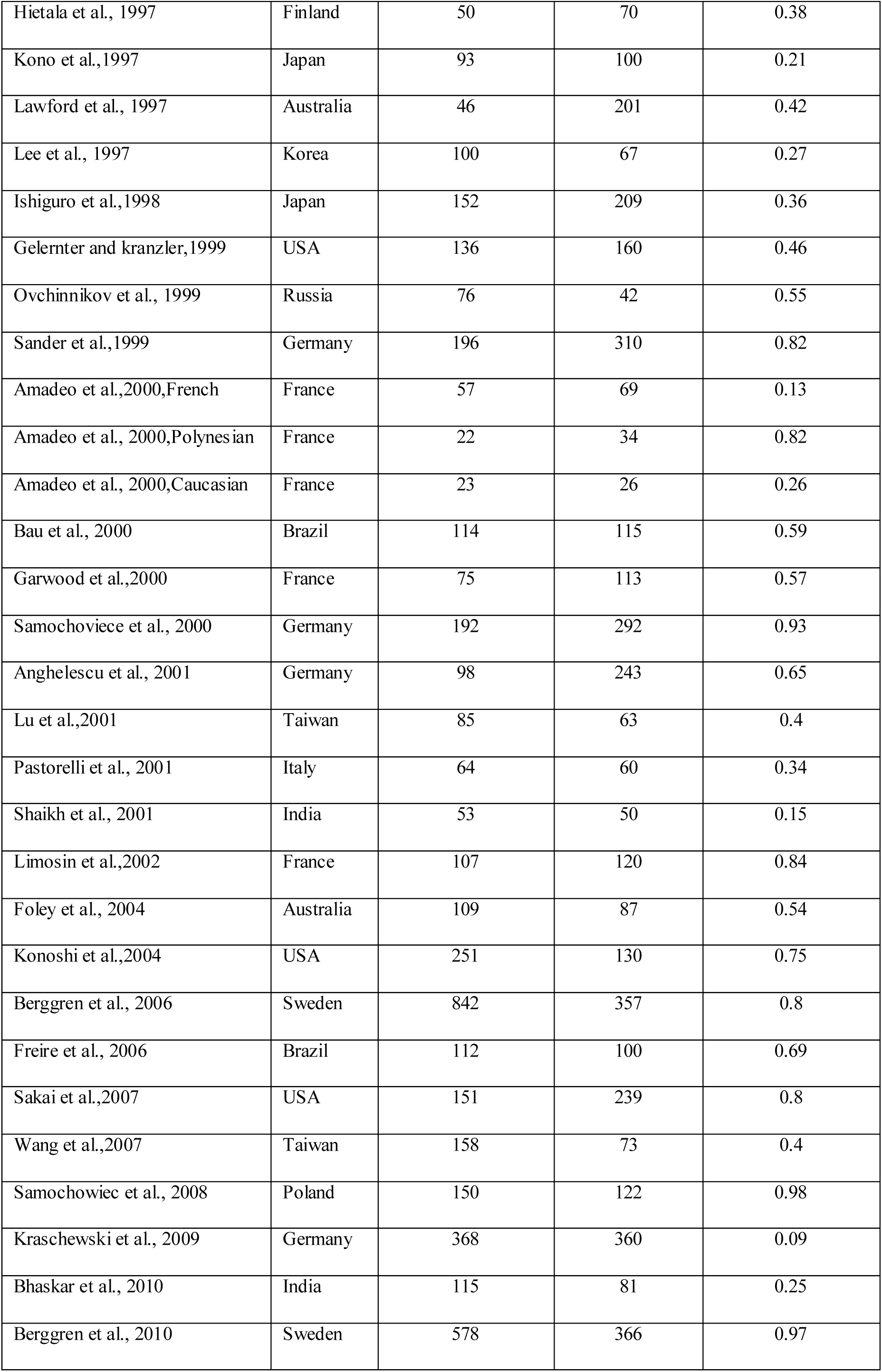
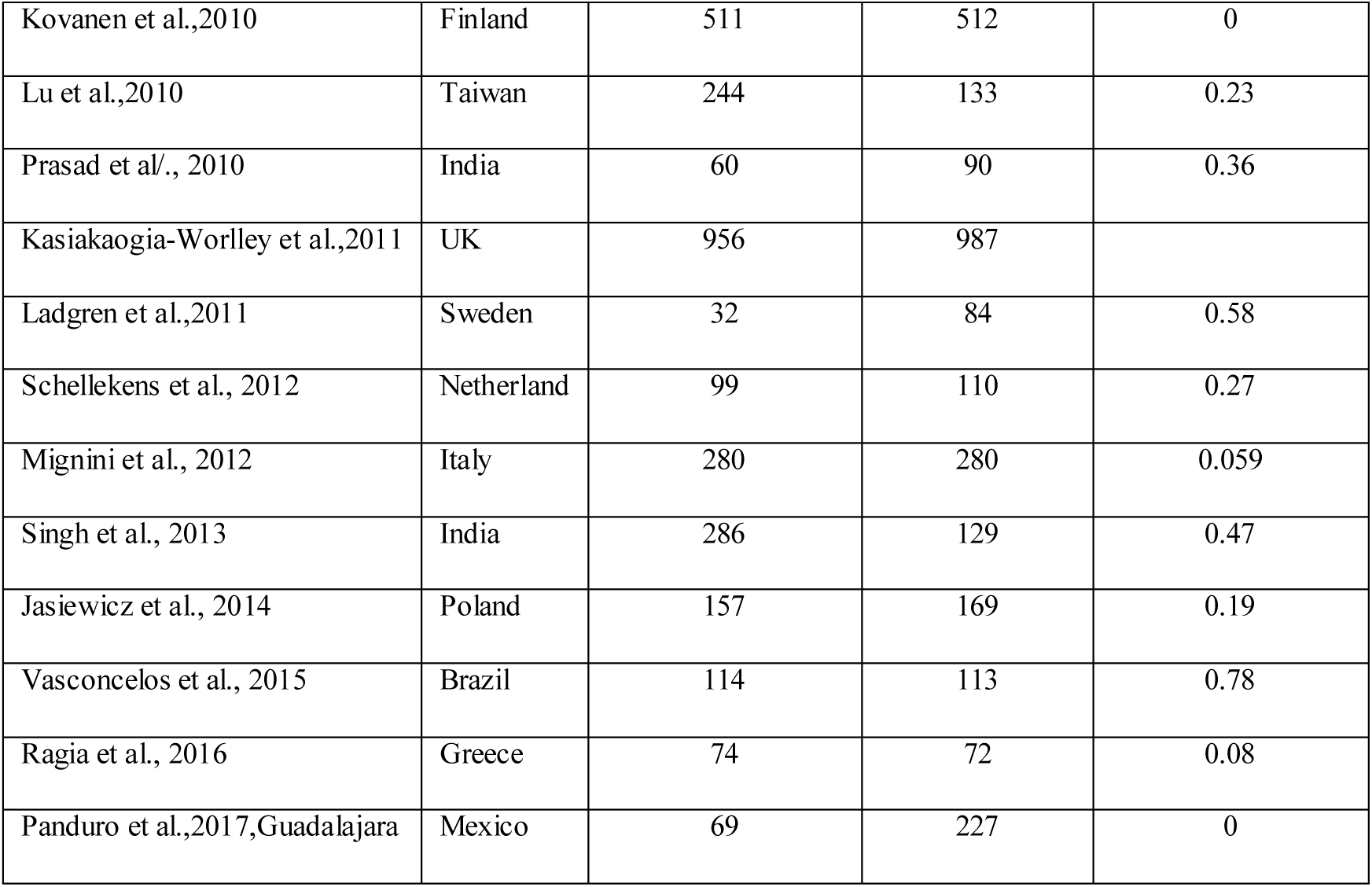
Details of included sixty nine studies.

| **Study** | **Country** | **Control number** | **Case number** | **p- value of HWE** |
| --- | --- | --- | --- | --- |
| Blum et al., 1990 | USA | 35 | 35 | 0.65 |
| Bolos et al., 1990 | USA | 127 | 40 | 0.02 |
| Blum et al., 1991 | USA | 43 | 96 | 0.55 |
| Comings et al., 1991 | USA | 108 | 104 | 0.19 |
| Gelernter et al., 1991 | USA | 68 | 44 | 0.81 |
| Parsian et al., 1991 | USA | 25 | 32 | 0.75 |
| Schwab et al., 1991 | Germany | 69 | 45 | 0.0007 |
| Cook et al., 1992 | Iowa | 20 | 20 | 0.42 |
| Goldman et al., 1992 | USA | 36 | 46 | 0.65 |
| Amadeo et al., 1993 | France | 43 | 49 | 0.56 |
| Arinami et al., 1993 | Japan | 35 | 78 | 0.17 |
| Geijer et al., 1994 | Sweden | 81 | 93 | 0.33 |
| Noble et al., 1994 | USA | 80 | 73 | 0.24 |
| Saurez et al., 1994 | USA | 89 | 88 | 0.95 |
| Neiswanger et al., 1995 | USA | 30 | 52 | 0.69 |
| Sanders et al.,1995 | Germany | 113 | 270 | 0.95 |
| Chen et al., 1996 | Taiwan | 41 | 73 | 0.14 |
| Heinz et al., 1996 | Germany | 113 | 97 | 0.95 |
| Lu et al.,1996,Han | Taiwan | 24 | 20 | 0.32 |
| Lu et al.,1996, Atyal | Taiwan | 21 | 21 | 0.15 |
| Lu et al.,1996,Ami | Taiwan | 20 | 20 | 0.06 |
| Chen et al.,1997,Atyal | Taiwan | 31 | 36 | 0.33 |
| Chen et al.,1997,Ami | Taiwan | 23 | 24 | 0.43 |
| Chen et al.,1997, Bunun | Taiwan | 58 | 58 | 0.47 |
| Chen et al., 1997, Paiwan | Taiwan | 35 | 35 | 0.54 |
| Chen et al.,1997, Han | Taiwan | 66 | 50 | 0.1 |
| Goldman et al.,1997 | USA | 161 | 276 | 0.26 |
| Hietala et al., 1997 | Finland | 50 | 70 | 0.38 |
| Kono et al.,1997 | Japan | 93 | 100 | 0.21 |
| Lawford et al., 1997 | Australia | 46 | 201 | 0.42 |
| Lee et al., 1997 | Korea | 100 | 67 | 0.27 |
| Ishiguro et al.,1998 | Japan | 152 | 209 | 0.36 |
| Gelernter and kranzler,1999 | USA | 136 | 160 | 0.46 |
| Ovchinnikov et al., 1999 | Russia | 76 | 42 | 0.55 |
| Sander et al.,1999 | Germany | 196 | 310 | 0.82 |
| Amadeo et al.,2000,French | France | 57 | 69 | 0.13 |
| Amadeo et al., 2000,Polynesian | France | 22 | 34 | 0.82 |
| Amadeo et al., 2000,Caucasian | France | 23 | 26 | 0.26 |
| Bau et al., 2000 | Brazil | 114 | 115 | 0.59 |
| Garwood et al.,2000 | France | 75 | 113 | 0.57 |
| Samochowiec et al., 2000 | Germany | 192 | 292 | 0.93 |
| Anghelescu et al., 2001 | Germany | 98 | 243 | 0.65 |
| Lu et al.,2001 | Taiwan | 85 | 63 | 0.4 |
| Pastorelli et al., 2001 | Italy | 64 | 60 | 0.34 |
| Shaikh et al., 2001 | India | 53 | 50 | 0.15 |
| Limosin et al.,2002 | France | 107 | 120 | 0.84 |
| Foley et al., 2004 | Australia | 109 | 87 | 0.54 |
| Konoshi et al.,2004 | USA | 251 | 130 | 0.75 |
| Berggren et al., 2006 | Sweden | 842 | 357 | 0.8 |
| Freire et al., 2006 | Brazil | 112 | 100 | 0.69 |
| Sakai et al.,2007 | USA | 151 | 239 | 0.8 |
| Wang et al.,2007 | Taiwan | 158 | 73 | 0.4 |
| Samochowiec et al., 2008 | Poland | 150 | 122 | 0.98 |
| Kraschewski et al., 2009 | Germany | 368 | 360 | 0.09 |
| Bhaskar et al., 2010 | India | 115 | 81 | 0.25 |
| Berggren et al., 2010 | Sweden | 578 | 366 | 0.97 |
| Kovanen et al.,2010 | Finland | 511 | 512 | 0 |
| Lu et al.,2010 | Taiwan | 244 | 133 | 0.23 |
| Prasad et al., 2010 | India | 60 | 90 | 0.36 |
| Kasiakaogia-Worley et al.,2011 | UK | 956 | 987 |  |
| Ladgren et al.,2011 | Sweden | 32 | 84 | 0.58 |
| Schellekens et al., 2012 | Netherland | 99 | 110 | 0.27 |
| Mignini et al., 2012 | Italy | 280 | 280 | 0.059 |
| Singh et al., 2013 | India | 286 | 129 | 0.47 |
| Jasiewicz et al., 2014 | Poland | 157 | 169 | 0.19 |
| Vasconcelos et al., 2015 | Brazil | 114 | 113 | 0.78 |
| Ragia et al., 2016 | Greece | 74 | 72 | 0.08 |
| Panduro et al.,2017,Guadalajara | Mexico | 69 | 227 | 0 |

**Table 2.** Summary estimates for the odds ratio (OR) in various allele/genotype contrasts, the significance level (p value) of heterogeneity test (Q test), and the I^2^ metric: overall analysis

| **Genetic Models** | **Fixed effect**<br>**OR (95% CI), p** | **Random effect**<br>**OR (95% CI), p** | **Heterogeneity**<br>**p-value** | **I<sup>2</sup> (%)** | **p-value**<br>**publication bias** |
| --- | --- | --- | --- | --- | --- |
| Allele Contrast (C vs TA) | 1.18 (1.12-1.24), <0.0001 | 1.22(1.13-1.31), <0.0001 | 0.0003 | 41.02 | 0.003 |
| Co-dominant (CT vs CC) | 1.26(1.17-1.36), <0.0001 | 1.27(1.15-1.40), <0.0001 | 0.005 | 33.2 | 0.19 |
| Homozygote (TT vs CC) | 1.35(1.18-1.55), <0.0001 | 1.39(1.19-1.61), <0.0001 | 0.27 | 8.95 | 0.42 |
| Dominant (TT+CT vs CC) | 1.29(1.20-1.39), <0.0001 | 1.29(1.18-1.43), <0.0001 | 0.002 | 36.57 | 0.18 |
| Recessive (TT vs CT+CC) | 1.21(1.08-1.36), 0.0006 | 1.26(1.11-1.43), 0.0004 | 0.32 | 6.45 | 0.06 |

Allele contrast meta-analysis revealed a modest significant association between AD and DRD2 gene T allele (T vs. C) with both fixed effects (OR= 1.18, 95% CI= 1.12-1.24, p<0.0001) and random effects model(OR=1.22, 95% CI=1.13-1.32, p<0.0001) (Table 2). Heterogeneity was less than 50% (I^2^=41.02), hence fixed effect mode was adopted. Results showed an increased risk of AD among mutant homozygote variants (TT vs.CC; homozygote model), with both fixed (OR= 1.35, 95%CI= 1.18-1.55; p= <0.0001) and random (OR= 1.39; 95%CI= 1.19-1.61; p<0.0001) effect models.

Mutant genotypes (TT+CT vs.CC; dominant model) showed positive significant association with AD using both fixed (OR= 1.29; 95%CI= 1.20-1.39; p<0.0001) and random (OR= 1.29; 95%CI= 1.18-1.453; p<0.0001) effect models (Table 2). Similarly the recessive genotypes model (TT vs. CT+CC) also showed significant association with AD with both fixed (OR= 1.21; 95%CI= 1.08-1.36; p= 0.0006) and random (OR= 1.26; 95%CI= 1.11-1.43; p= 0.0004) effect models (Table 2).

### Sub-group Analysis

Authors also performed ethnicity based sub-group analysis. Out of 69 studies, 20 studies were from Asia and 49 studies were from Caucasian population. In Asian population (number of studies= 20; 1410/1700 cases/controls), meta-analysis using all five genetic modes did not show any significant association (allele contrast: OR= 1.23, 95% CI= 1.11-1.37, p=0.87; dominant model: OR = 1.31, 95% CI= 1.13-153, p=0.71) (Figure 3). Results of Caucasian studies (number of studies= 49; 7715/7423 cases/controls) meta-analysis using a five genetic modes indicated significant association with both fixed and random effects model, but fixed effect mode was adopted due to less heterogeneity (allele contrast: OR= 1.23, 95% CI= 1.23-1.35, p=0.00; dominant model: OR = 1.31, 95% CI= 1.16-147, p=0.00) (Figure 4).

**Figure 2.**
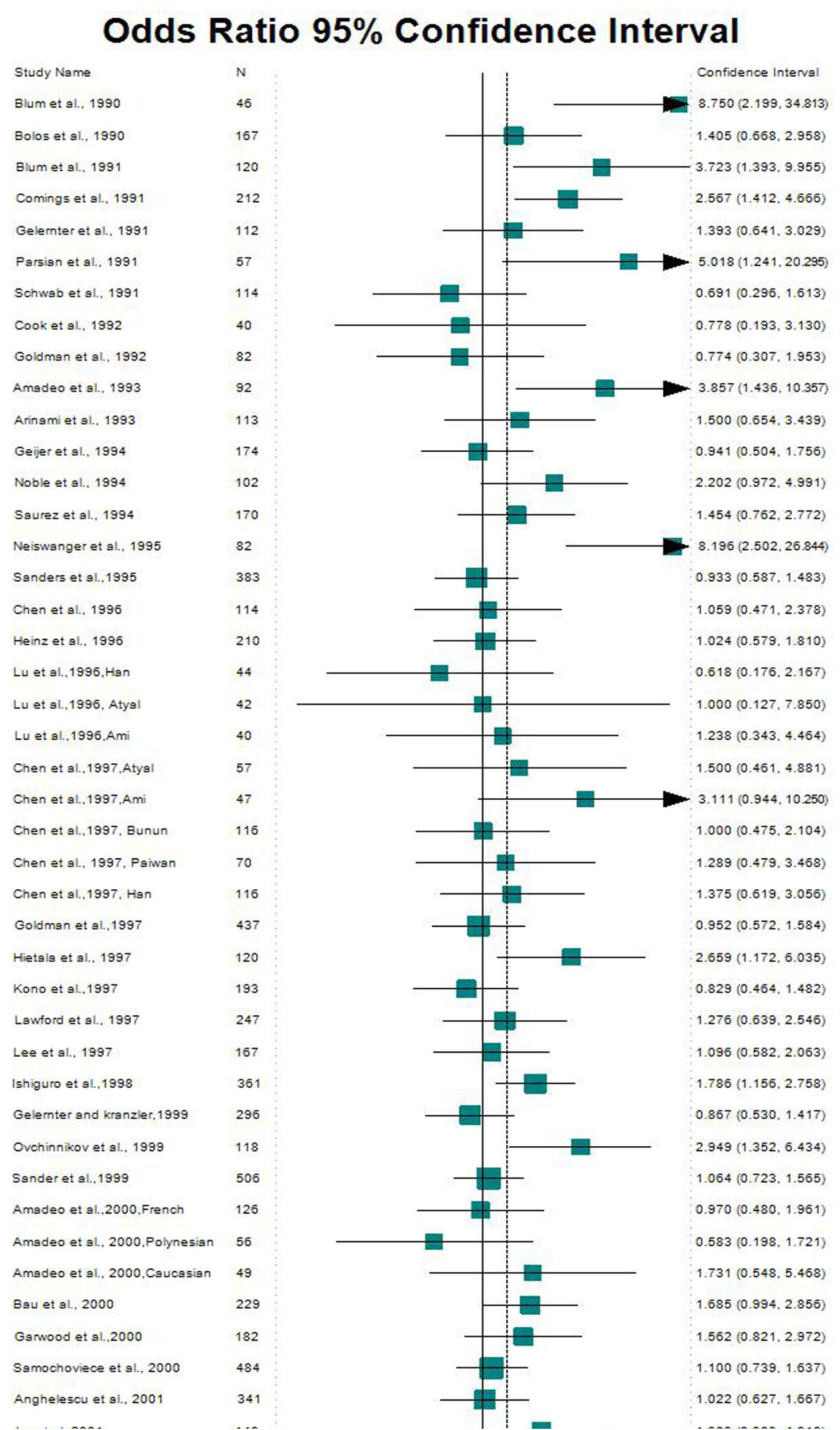
Forest Plot of dominant model of 69 studies.

**Figure 3.**
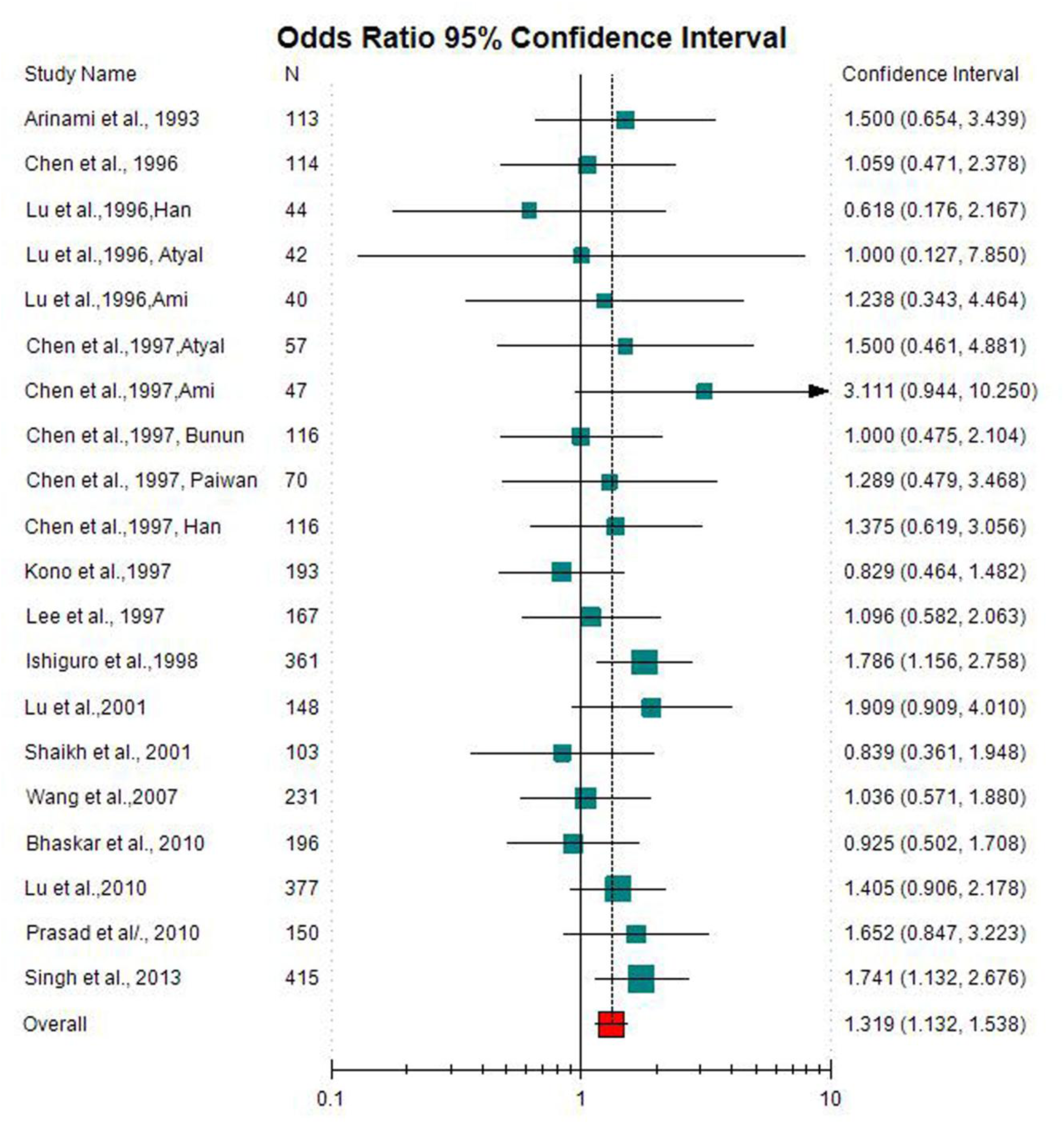
Forest plot of dominant model of Asian studies.

**Figure 4.**
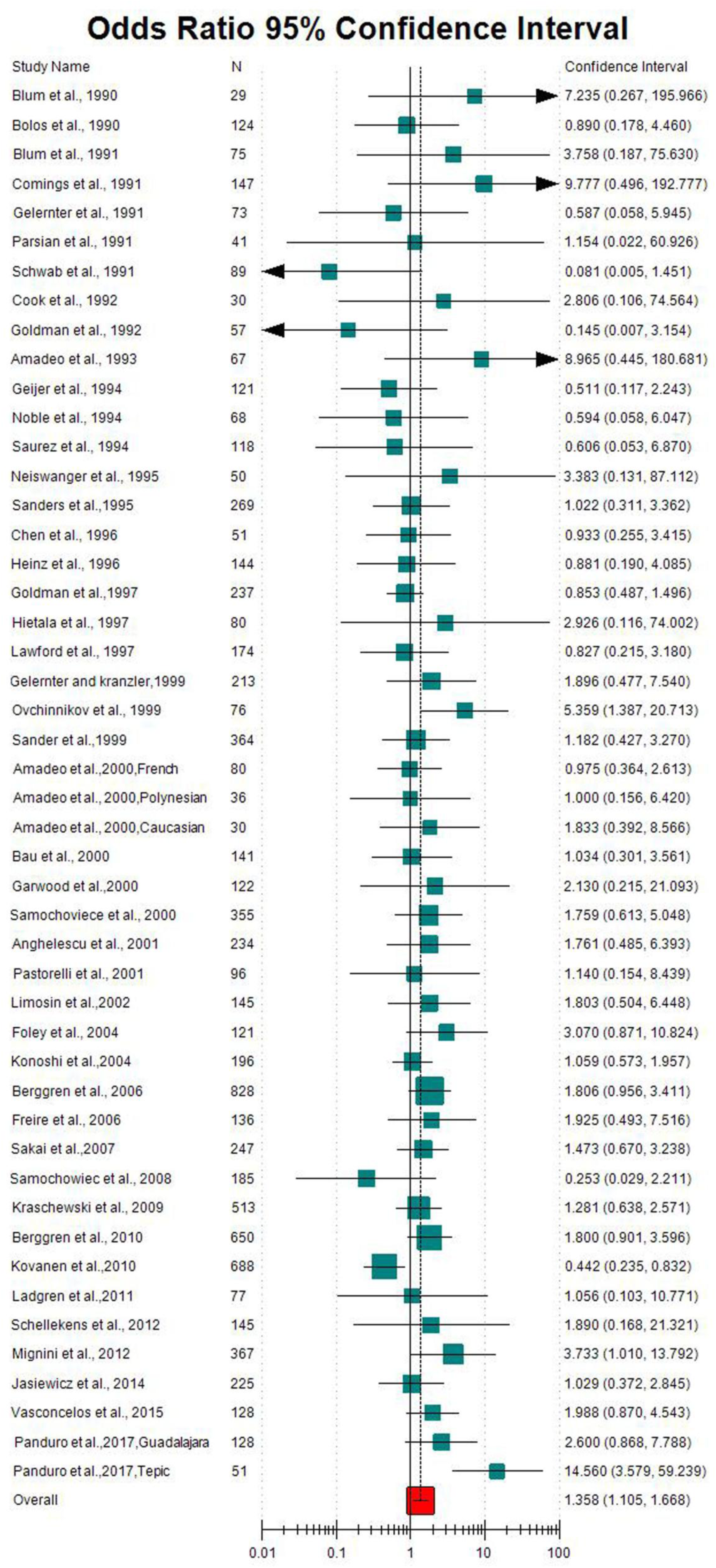
Forest plot of homozygote model of Caucasian studies.

**Figure 5.**
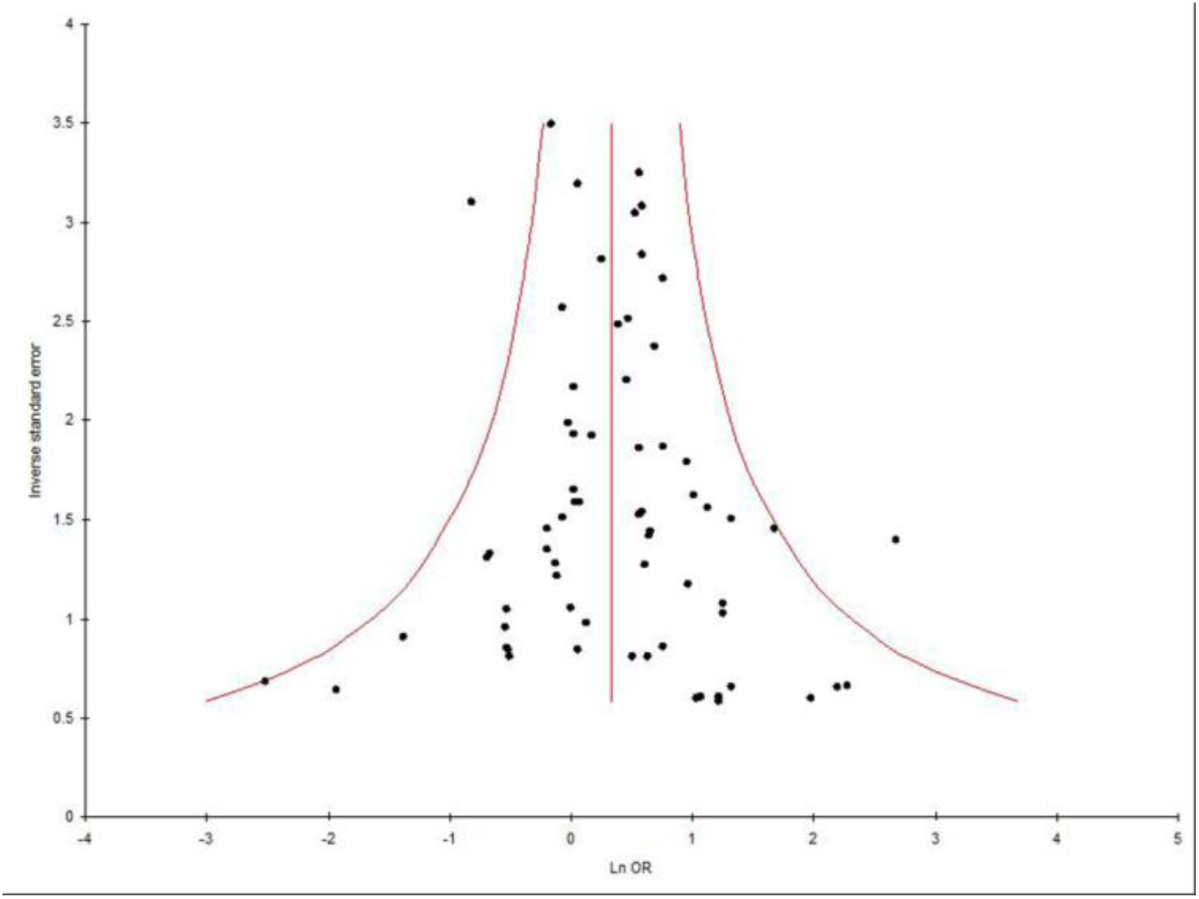
Funnel plot of allele contrast model of 69 studies.

### Sensitivity analysis

In allele contrast meta-analysis, sensitivity analysis performed by exclusion of five studies in which control population was not in HWE. Result of meta-analysis by exclusion of these five studies not in HWE (Bolos et al.,1990; Schwab et al.,1991; Kovanen et al.,2010; Panduro et al., 2017-Guadalaraja; Panduro et al., 2017-Tepic) showed that odds ratio was not increased. In addition, the influence of each study on the pooled OR was examined by repeating the meta-analysis while omitting each study one at a time. The results suggested that no individual study significantly affected the pooled ORs.

### Publication bias

Begg’s funnel plot and Egger’s test were performed to assess the publication bias. The shape of funnel plots did not reveal any evidence of obvious asymmetry in all genetic models except allele contrast (Figure). Egger’s test was used to provide statistical evidence and p values of Egger’s tests were more than 0.05 (p=0.003 for T *vs* C; p=0.42 for TT *vs* CC; p=0.19 for CT *vs* CC; p=0.18 for TT+CT *vs* CC; and p=0.06 for TT *vs* CT+CC). The results did not show any evidence of publication bias except in allele contrast meta-analysis of total studies. The detailed data were given in table 2.

## Discussion

It is well recognized that there is individual susceptibility to AD even within the same environmental exposure. Main factor, including polymorphisms of genes involved in reward cascade pathway may have accounted for this difference. A1 allele of the DRD2 Taq1A polymorphism was reported to be associated with altered/reduced D2 receptor expression in the brain and may confer susceptibility to AD (Pohjalainen et al., 1998).

Four meta-analyses are published and demonstrated a moderate effect of the Taq1A polymorphism on AD (Munafo et al. 2007; Le Foll et al. 2009; Smith et al., 2008; Wang et al., 2013). Present meta-analysis included the largest number of studies so far investigating the association between the Taq1A polymorphism and AD. Sixty-nine eligible studies published up to 2017 were considered, including 9,125 cases and 9,123 healthy controls. Our results provide strong evidence of the association between the Taq1A polymorphism and AD, especially in the European population. Heterogeneity and publication bias are not present and sensitivity analysis showed that the results from both allelic and genotypic meta-analyses are stable and not influenced by any individual study.

Meta-analysis offers a powerful method to synthesize information of independent studies with similar target (Ioannidis et al., 2002). Several meta-analysis are published, which evaluated effects of polymorphism in susceptibility of diseases/disorders-down syndrome (Rai,2011; Rai et al., 2017; Rai and Kumar, 2018), cleft lip and palate (Rai,2014, 2017), Glucose-6-phosphate dehydrogenase deficiency (Kumar et al.,2016), male infertility (Rai and Kumar,2017),schizophrenia (Yadav et al., 2015; Rai et al., 2017), obsessive compulsive disorder (Kumar and Rai,2019), depression (Rai,2014,2017), epilepsy (Rai and Kumar,2018),Alzheimers disease (Rai, 2016), esophageal cancer (Kumar and Rai, 2018), prostate cancer (Yadav et al.,2016), breast cancer (Kumar et al., 2015; Rai et al.,2017), digestive tract cancer (Yadav et al.,2018), ovary cancer (Rai, 2016), endometrial cancer (Kumar et al., 2018), uterine leiomyioma (Kumar and Rai, 2018), MTHFR gene frequency (Yadav et al.,2018), and MTRR gene frequency (Yadav et al.,2019).

Along with strengths, there were also few limitations in current meta-analysis like-(i) crude OR was used in meta-analysis, (ii) other risk factors among the subjects in the available studies, such as smoking status, diet etc were not considered and (iii) only single gene polymorphism was considered and gene -gene or gene-environment interactions were not considered.

In conclusion, pooled analysis of data from 69 separate populations indicates that the DRD2 Taq1A polymorphism is associated with significant risk of AD. Results of subgroup analyses showrd that TaqIA polymorphism is not risk factor for AD in Asian population, but this polymorphism is provides susceptibility for AD in Caucasian population. Future large-scale, population-based association studies are now need of the hour to investigate potential gene–gene and gene–environment interactions involving the DRD2 Taq1A polymorphism in determining risk of AD.

## Data Availability

All data available in the manuscript.

